# Developing a taxonomy of care coordination for people living with rare conditions: A qualitative study

**DOI:** 10.1101/2021.11.16.21266387

**Authors:** Holly Walton, Amy Simpson, Angus I.G. Ramsay, Emma Hudson, Amy Hunter, Jennifer Jones, Pei Li Ng, Kerry Leeson-Beevers, Lara Bloom, Joe Kai, Larissa Kerecuk, Maria Kokocinska, Alastair G Sutcliffe, Stephen Morris, Naomi J Fulop

## Abstract

**Background:** Improving care coordination is particularly important for individuals with rare conditions (who may experience multiple inputs into their care, across different providers and settings). To develop and evaluate strategies to potentially improve care coordination, it is necessary to develop a method for organising different ways of coordinating care for rare conditions. Developing a taxonomy would help to describe different ways of coordinating care and in turn facilitate development and evaluation of pre-existing and new models of care coordination for rare conditions. To the authors’ knowledge, no studies have previously developed taxonomies of care coordination for rare conditions. This research aimed to develop and refine a care coordination taxonomy for people with rare conditions.

**Methods:** This study had a qualitative design and was conducted in the United Kingdom. To develop a taxonomy, six stages of taxonomy development were followed. We conducted interviews (n=30 health care professionals/charity representatives/commissioners) and focus groups (n=4 focus groups, 22 patients/carers with rare/ultra-rare/undiagnosed conditions). Interviews and focus groups were audio-recorded with consent, and professionally transcribed. Findings were analysed using thematic analysis. Themes were used to develop a taxonomy, and to identify which types of coordination may work best in which situations. To refine the taxonomy, we conducted two workshops (n=12 patients and carers group; n=15 professional stakeholder group).

**Results:** Our taxonomy has six domains, each with different options. The six domains are: 1) Ways of organising care (local, hybrid, national), 2) Ways of organising professionals involved in care (collaboration between many or all professionals, collaboration between some professionals, a lack of collaborative approach), 3) Responsibility for coordination (administrative support, formal roles and responsibilities, supportive roles and no responsibility), 4) How often appointments and coordination take place (regular, on demand, hybrid), 5) Access to records (full or filtered access), and 6) Mode of care coordination (face-to-face, digital, telephone).

**Conclusions:** Findings indicate that there are different ways of coordinating care across the six domains outlined in our taxonomy. This may help to facilitate the development and evaluation of existing and new models of care coordination for people living with rare conditions.

## Introduction

The complexity of the organisation and delivery of health care has been further complicated in recent years, due to the need to manage a higher demand for health care and the introduction of new technologies, increased availability of treatments and provision of care across many settings [1]. These changes and demand on health care may make it difficult for health care organisations to manage care [1], for providers to deliver care, and may make it more burdensome for patients to receive and engage with their care [2,3,4]. One potential solution to these challenges is to consider and further develop models of care, such as care coordination [1]. Enhanced coordination is particularly beneficial for those with complex conditions such as chronic [2,4] and rare conditions [3,5,6,7,8]. Rare, ultra-rare and undiagnosed conditions are often complex, and affect multiple body systems and a person’s mental and physical health [9,10]. Rare conditions require care from multiple sectors, and health care professionals. For example, patients may be seen by primary care, secondary care, tertiary and quaternary care providers. Whilst individually rare conditions only affect a few individuals (each condition affects up to five in 10,000 people) [6,9], collectively the 6000-8000 rare and ultra-rare conditions together with undiagnosed conditions affect a significant proportion of individuals worldwide [11].

Previous research has indicated that a lack of coordination has many negative impacts on patients and carers living with rare conditions, including on their physical health, finances, psychological wellbeing, and social aspects of their lives [12]. A recent scoping review of 154 reviews of common and rare chronic conditions, together with focus groups with patients, carers and health care professionals, has defined care coordination for rare conditions [13]. Findings indicated that coordination for rare conditions has many components and that there are many different options for how care can be coordinated [13]. Coordination was defined as everyone involved in a person’s care working together across multiple aspects of care to avoid duplication and achieve shared outcomes. Coordination would need to be lifelong and involve all parts of the health and social care system (including different services, settings, multiple conditions, and transition between services). It has been argued that coordination should be family-centred, evidence-based and ensure equal access for all [13]. The review highlighted many different components of coordination, including components that need to be coordinated (e.g., assessment), components that inform how to coordinate care (e.g., someone to take responsibility), components that have multiple roles (e.g., planning) and components that contextualise coordination (e.g., evidence-based practice) [13].

In order to better understand and potentially improve care coordination, it is necessary to identify and describe the different ways in which care can be coordinated for rare conditions. One way to facilitate the organisation of care coordination is to develop a taxonomy of care coordination for rare conditions. Taxonomies are systems used to organise complex concepts into common conceptual domains and dimensions based on similarities [14,15]. Developing a taxonomy of care coordination for rare conditions would help to describe different ways of coordinating care. This in turn can facilitate the development and evaluation of pre-existing and new models of care coordination.

Taxonomies aim to provide clear definitions [15], and have been previously used to organise complex health care concepts including taxonomies of integrated care [16], health care [17], behaviour change [18], and the burden of treatment for patients with chronic conditions [19]. For example, the burden of treatment taxonomy included tasks that the health care system imposes on patients, factors worsening the burden of treatment and consequences of burden from the patients’ point of view [19].

To the authors’ knowledge, no previous studies have attempted to develop a taxonomy of coordination of care specifically for rare or chronic conditions. This study aimed to develop and refine a taxonomy of care coordination for people living with rare conditions, based on learning from the UK healthcare context and the National Health Service. Whilst there are many different rare conditions and the care needs for each of these may differ slightly, it is necessary to develop a taxonomy that can be used to outline the different options and then these options can be adapted and applied to suit different contexts and different rare, ultra-rare and undiagnosed conditions. We present our findings on what types of coordination may be appropriate in different situations and the development of hypothetical models of care coordination separately [20].

## Methods

This study is part of a larger mixed-methods research project which aims to explore coordination of care for people living with rare conditions [21]. This study builds on previous aspects of this study [12, 13.

A summary of the methods used in this study are provided below (see Table 1 for a detailed description).

**Table 1.** Detailed description of the methods used in this study.

|  | Development of taxonomy (interviews and focus groups) | Refinement of taxonomy (workshops) |
| --- | --- | --- |
| **Design** | Previously both quantitative [22, 23] and qualitative [15, 19,24,25] approaches have been used to develop taxonomies.<br>Our study used qualitative methods (interviews, focus groups and workshops) and was conducted in a two-stage process (see Figure 1). First, interviews and focus groups were conducted to develop an initial taxonomy. Workshops were then conducted to refine the proposed taxonomy.<br>Qualitative methods were used as they allowed for more in-depth understanding of coordination which is a multifaceted concept [24,25]. Additionally, qualitative methods enabled us to involve stakeholders with the most experience of care coordination, which is particularly important in health care service research [26]. By understanding patients', carers' and health care professionals' views on the organisation of coordination of care for rare conditions it may be possible to improve health care services, and optimise the patient experience. |  |
| **Setting** | UK based study<br>Coordination across the NHS, social care and third sector (with a primary focus on health care). |  |
| **Sample** | **Eligibility criteria**<br>To participate in our study, participants needed to be:<br>18 or over (Children were not included due to ethical issues recruiting participants under 18).<br>Patients with rare, ultra-rare or undiagnosed conditions, carers/parents of children or adults with a rare, ultra-rare or undiagnosed condition, health care professionals, charity representatives or commissioners.<br>**Recruitment methods**<br>Participants were recruited using a range of methods, including:<br>Email invitation<br>Adverts on social media<br>Voluntary sector study advertisement<br>Adverts through our partnership with four NHS sites<br>**Sampling criteria**<br>To ensure that different models of coordinated care and a wide range of experience and expertise were captured, we purposively sampled using the following characteristics.<br>For health care professionals/commissioners/charity representatives – area of the UK, job role, experience with different types of care coordination<br>For patients/carers – area of UK, condition, role, age, experience with different types of care coordination. |  |
| **Measures** | To develop the taxonomy, we developed two topic guides: i) interview topic guide and ii) focus group topic guide.<br>Questions focused on a range of topics including stakeholders' experiences of coordinated care, implications of coordinated care, preferences for aspects of care coordination (ways of coordinating care, format, access, frequency, location, information sharing, transition), benefits and challenges and factors that help or hinder coordination.<br>Feedback on the topic guide was sought from the CONCORD public and patient involvement advisory group prior to data collection. | We developed one topic guide for the workshops.<br>The topic guide was based around the six categories identified in the taxonomy and included prompts on whether we had missed anything, whether findings seemed appropriate based on participants' experiences, appropriateness of options in light of the COVID-19 pandemic and recommendations to improve each of the six categories. |
| **Procedure – Recruitment and ethics** | Potential participants contacted the researcher (HW) via email or telephone and were given participant information sheets.<br>Potential participants were asked to provide responses to eligibility questions when registering their interest.<br>For professionals, these included: their occupation, speciality and geographical region.<br>For patients or carers, these included whether they receive coordinated care (specialist service and who coordinates), whether they have a diagnosis, are a patient/carer, their age range, ethnicity and geographical region.<br>Selected individuals were asked to complete two written consent forms prior to taking part in the interviews, focus groups or workshops. Participants who took part virtually or via telephone were asked to return written consent forms in advance.<br>Participants were informed that their data would be kept confidential, fully anonymised and that they could withdraw at any time without reason. Focus group participants were informed that any data collected up until the point of withdrawal would be kept due to difficulties removing individual participants from focus group data. We took steps to ensure that quotes from the |  |
|  | participant who withdrew from the study were not included in publications. |  |
| **Procedure – Data collection** | - One researcher (HW) conducted the interviews<br>- Interviews took place by phone (n=27) or face-to-face (n=3), depending on participants' preferences.<br>- The interviews lasted approximately one hour (range: 44 to 74 minutes).<br>- Two researchers (HW and AS) conducted the four focus groups (one researcher facilitated, and one researcher took notes) [27]. A third researcher observed one of the focus groups (EH). Two focus groups were face-to-face in two cities in the UK, and two were conducted using Skype for Business. Focus groups were up to three hours in length (including a break) (range: 149-154 minutes).<br>- Interviews and focus groups were digitally recorded using an encrypted dictaphone (with consent from participants) and professionally transcribed.<br>- Transcripts were checked for accuracy and fully anonymised (including names, places and due to their rareness – the names of specific conditions).<br>- Data were stored in the university's secure data environment and coded using NVivo 12 [28]. | - Workshop participants were sent a 15-minute video prior to the workshop which outlined the findings of the taxonomy.<br>- Participants were split into three breakout groups.<br>- Each breakout group had one facilitator (HW, EH, AlGR) and one note taker (JJ, SM, AH).<br>- After the breakout groups, participants reconvened in the main group and received feedback from each group on their discussions.<br>- Workshops were recorded using an encrypted Dictaphone.<br>- Notes were checked for thoroughness and summarised prior to being sent to a graphic facilitator (New Possibilities) to create a graphical representation of the findings. |
| **Analysis** | - Thematic analysis was used to analyse interview and focus group data. In line with recommendations for taxonomy development [15].<br>- Inductive coding was used to develop an initial coding frame [29]. Six interview transcripts were coded inductively by two researchers (HW/AS). A coding framework was developed and agreed. The framework included codes on aspects of care coordination (types, who is involved, mode, information sharing, where, frequency, transition between services, methods of access) and qualifier codes (preferences, benefits and challenges, barriers and facilitators, factors influencing coordination).<br>- The framework was used to code all interview and focus group transcripts (HW). A second researcher (AS) coded six interviews and one focus group transcript (20% of the data). Coding was discussed and any discrepancies (e.g. on how codes were used/what codes meant/when to use codes) were agreed.<br>- Findings were then grouped into themes and sub-themes using thematic analysis [30]. Given the large amount of data, this was done in two stages: 1) development of themes and sub-themes for the data on aspects of coordination (to develop initial taxonomy options), 2) development of themes and sub-themes for the data on qualifying codes (to develop models) (described in [20]).<br>- Five themes were developed (ways of organising care, ways of organising teams, responsibilities for coordination, access to coordination and mode of coordination).<br>- Themes and sub-themes were discussed by co-authors and used to develop a taxonomy.<br>- Once themes and sub-themes had been developed Six stages of taxonomy development [31] were followed:<br>1) **Identify the meta-characteristic** that will inform the choice of characteristics in the taxonomy.<br>2) **Identify ending conditions** (requirements that the taxonomy needs to meet to be finalised).<br>3) **Choose approach.** We used an empirical-conceptual approach. We based the taxonomy on our findings from interviews and focus groups and earlier CONCORD findings<br>4) **Identify a subset of objects to classify**, using findings from the interviews and focus groups.<br>5) **Identify common characteristics** (similarities and differences will be identified to identify common characteristics and discriminatory characteristics for coordinated care).<br>6) **Group the characteristics** using a manual process [14].<br>- We reviewed our findings in relation to the CONCORD scoping review [13] and preliminary survey findings [21] to ensure that there were no major options missing from our taxonomy. | - Workshop notes were coded and grouped into themes surrounding their experiences of the model of coordination, benefits and challenges of the model of coordination, factors influencing coordination, missing aspects and impact of COVID-19.<br>- Feedback on aspects that were missing in the taxonomy were used to refine and finalise the taxonomy.<br>- Findings highlighted key aspects to be clarified within the taxonomy, including: the need to emphasise that care is not just medical (also includes social and educational aspects), and that care is lifelong.<br>- Findings also highlighted the need to separate out collaborations that include patients/carers from collaborations between professionals. The need for third sector involvement in collaboration where appropriate; the need to emphasise the role that charities and patients/carers play in care coordination; a hybrid model of frequency and the need to clarify aspects of the mode domain.<br>- The taxonomy was amended in line with this feedback. |
|  | - Additionally, the wider CONCORD team and CONCORD public patient involvement advisory group reviewed the taxonomy findings and provided feedback on the taxonomy prior to the workshop. |  |

### Design

A two-stage study using qualitative methods was conducted (see Figure 1). In stage one, interviews and focus groups were conducted to develop a taxonomy. In stage two, workshops were conducted to refine the taxonomy.

**Figure 1.**
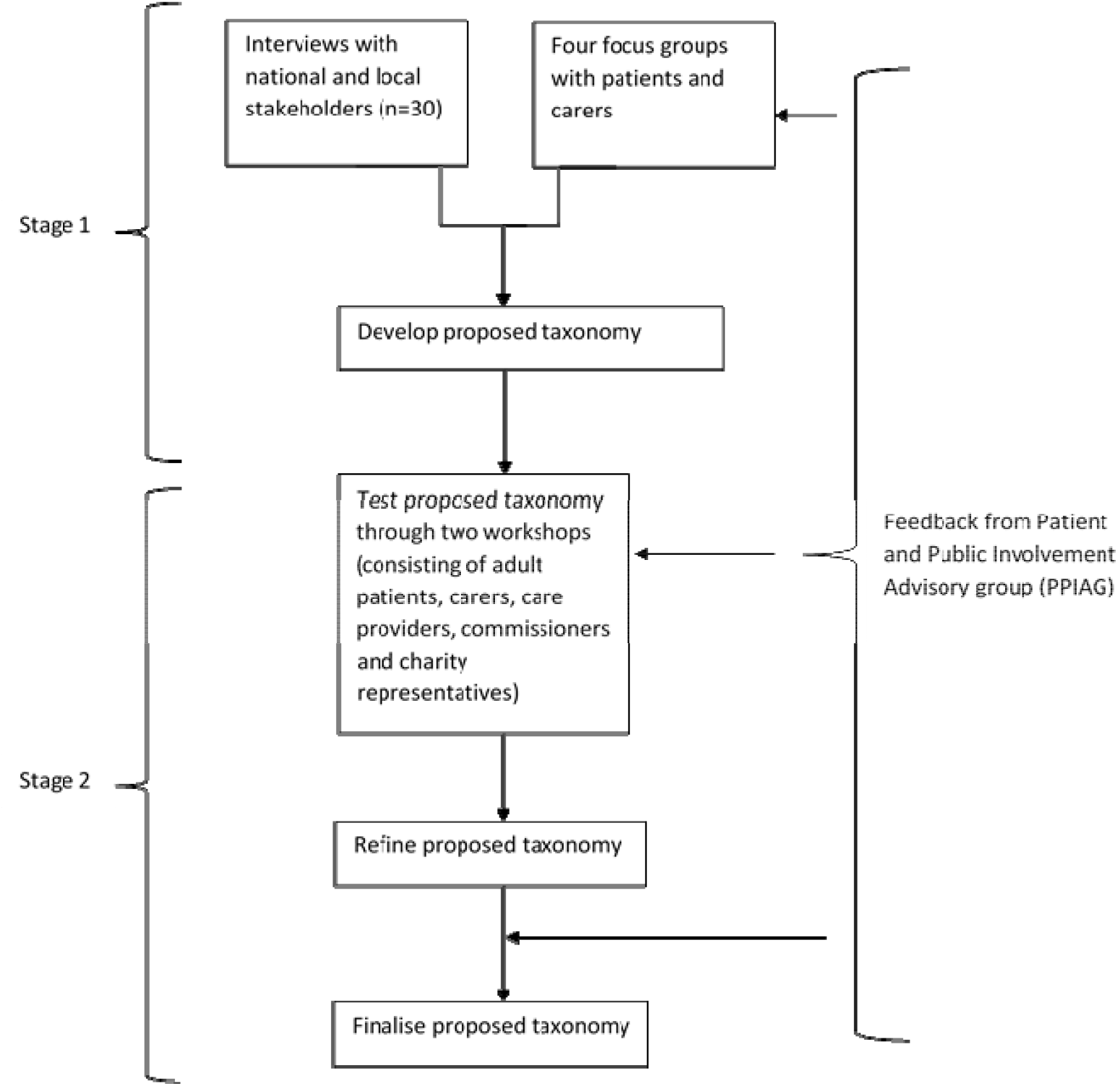
An overview of the two stages involved in this research.

### Setting

Our study explored care coordination in the UK, across different sectors; with a focus on health care and the National Health Service.

### Sample

Participants were recruited purposively using a range of methods including advertisements throughout the voluntary sector and advertisements through our partnership with four NHS sites. We recruited a range of individuals with experience of rare conditions, including patients, parents/carers, and professionals (health care professionals, charity representatives and commissioners). Participants took part in interviews (n=30 professionals), focus groups (four groups of 6-8 patients/carers) [27] and workshops (one patient/carer, and one professional; approximately 15 participants in each workshop). For patients and carers, we selected participants based on the area of the UK they lived in, their condition, role, age, and experience with different types of care coordination. For healthcare professionals, we selected participants based on the area of the UK they worked in, their job role and experience of different types of coordination. Eligibility criteria, recruitment methods and sampling criteria are provided in Table 1.

### Procedure

Study adverts included details of the researcher’s contact details and asked individuals to get in touch via email or telephone. Interested individuals contacted the researcher and were provided with an information sheet. Participants were asked to sign consent forms in advance of the interviews, focus groups and workshops. Participants were informed that their data would be kept confidential, fully anonymised and that they could withdraw without reason.

To develop the taxonomy, one researcher (HW) conducted interviews with health care professionals, commissioners and charity representatives. Two researchers (HW/AS), with a third observer (EH) conducted four focus groups with patients and carers (two face-to-face and two virtual). Interviews and focus groups were recorded with consent, professionally transcribed, checked for accuracy and fully anonymised. To develop the taxonomy and identify different ways of coordinating care, we used an interview topic guide and a focus group topic guide. See Table 1 for further details.

To refine the taxonomy, we held workshops with patients, carers, health care professionals, charity representatives and commissioners. We developed a workshop topic guide to determine if the findings were appropriate and comprehensive. Workshop participants were sent a video outlining the findings prior to the workshops. Participants were split into three breakout groups and asked to discuss the findings for each of the six taxonomy domains. Workshops were recorded and notes were taken. Notes from the workshop were also used to produce a graphical representation of findings.

For a more detailed description of our procedure, please see Table 1.

### Analysis

Thematic analysis was used to analyse interview and focus group data. In line with recommendations for taxonomy development [15]. Inductive coding was used to develop the coding framework [29]. Two researchers (HW/AS) initially coded six interview transcripts to develop and agree a coding frame. The coding framework was then used to code all interview and focus group transcripts by one researcher (HW). A second researcher (AS) also coded 20% of the data. Findings were discussed and agreed. Findings were then grouped into themes and sub-themes. Given the large amount of data, this was done in two stages: 1) development of themes and sub-themes for the data on aspects of coordination (to develop initial taxonomy options), 2) development of themes and sub-themes for findings relating to which models of care coordination work in different situations (described in [20]). Themes were discussed by co-authors and used to develop a taxonomy. Findings were also used to explore which models of care coordination may be appropriate in different situations and to develop hypothetical models of care coordination (these findings are described elsewhere in [20]).

Once themes and sub-themes had been developed, we followed six stages of taxonomy development to develop the proposed taxonomy [32]: 1) Identify the meta-characteristic (focus of the taxonomy), 2) Identify ending conditions, 3) Choose approach, 4) Identify a subset of objects to classify, 5) Identify common characteristics and 6) Group the characteristics [14]. Table 2 outlines how we applied these six steps.

**Table 2.** Description of how we applied Nickerson et al’s [31] taxonomy development criteria.

| Step | Our process |
| --- | --- |
| 1) Identify meta-characteristic | Meta characteristic = different ways in which care can be coordinated for rare conditions |
| 2) Identify ending conditions | Our ending conditions:<br>1) Not merging or splitting any objects in the last iteration<br>2) Having at least one object (type of coordinated care) under every characteristic of every dimension<br>3) Not adding any new dimensions or characteristics in last iteration<br>4) Uniqueness of dimensions, characteristics and cells |
| 3) Decide on approach | We used an empirical-conceptual approach. We based the taxonomy on our findings from interviews and focus groups and earlier CONCORD findings |
| 4) Use a subset of objects to classify | We used themes and sub-themes from the interviews and focus groups as objects to classify. The sub-themes outline types of coordination that can be used as objects (e.g., nationally commissioned services and condition-specific clinics). List of 'objects' (example ways of coordinating care) were identified from themes and sub-themes. |
| 5) Identify common characteristic | Similarities and differences were identified to identify common characteristics and discriminatory characteristics. These were identified through the summaries of themes and sub-themes |
| 6) Group characteristics using a manual or graphical process | We used a manual process to group characteristics into domains to form the first draft of the taxonomy |

To refine the taxonomy, we coded workshop notes and grouped them into themes (see Appendix 3 for a visual representation of workshop findings). Findings were used to amend the taxonomy. See Table 1 for further details.

## Results

### Participant characteristics

This study included 77 different participants (two participants took part in both a workshop and interview/focus group). Participants included patients, carers, health care professionals, commissioners and charity representatives. Overall, data from 52 participants (30 individual interviews, 22 focus group participants) informed the development of the taxonomy. Data from 27 workshop participants informed the refinement of the taxonomy. Demographic characteristics are shown in Table 3.

**Table 3.** Demographic characteristics of participants

|  | Development of taxonomy (n=52) |  | Refinement of taxonomy (n=27) |  | Total |
| --- | --- | --- | --- | --- | --- |
|  | Interviews | Focus groups | Patient and carer workshop | Professional workshop |  |
| **Number of participants** | 30 | 22<sup>a</sup> | 12 | 15 | **79 (77 different people<sup>b</sup>)** |
| **Type of participant** |  |  |  |  |  |
| Patients | N/A | 16 | 5 | N/A | **21** |
| Parents/carers of children aged < 18 years | N/A | 5 | 4 | N/A | **9** |
| Parents/carers (e.g. spouses) of adults aged ≥ 18 years | N/A | 1 | 3 | N/A | **4** |
| Health care professionals<sup>c, h</sup> | 15 | N/A | N/A | 2 | **17** |
| Health care professionals employed by charity | 2 | N/A | N/A | 2 | **4** |
| Charity representatives<sup>d, h</sup> | 5 | N/A | N/A | 8 | **13** |
| Commissioners | 3 | N/A | N/A | 3 | **6** |
| Multiple professional roles<sup>e</sup> | 5 | N/A | N/A | N/A | **5** |
| **Age (years)** |  |  |  |  |  |
| 18-25 | N/A | 2 | 0 | N/A | **2** |
| 26-59 | N/A | 16 | 10 | N/A | **26** |
| ≥ 60 | N/A | 4 | 2 | N/A | **6** |
| **Diagnosis<sup>i</sup>** |  |  |  |  |  |
| Rare/ultra-rare condition(s) | N/A | 22 | 12 | N/A | **34** |
| **Attend specialised service<sup>f</sup>** |  |  |  |  |  |
| Yes | N/A | 14 | 6 | N/A | **20** |
| No | N/A | 7 | 4 | N/A | **11** |
| Not sure | N/A | 1 | 2 | N/A | **3** |
| **Locations represented** |  |  |  |  |  |
| National role (UK) | 2 | 0 | 0 | 8 | **10** |
| National role (England and Wales) | 1 | 0 | 0 | 1 | **2** |
| National role (England) | 5 | 0 | 0 | 3 | **8** |
| Scotland | 1 | 0 | 1 | 0 | **2** |
| Wales | 1 | 1 | 0 | 0 | **2** |
| East of England | 1 | 2 | 1 | 1<sup>g</sup> | **5** |
| London | 4 | 7 | 0 | 0 | **11** |
| Yorkshire and the Humber | 1 | 2 | 0 | 0 | **3** |
| North East of England | 1 | 2 | 0 | 0 | **3** |
| North of England | 1 | 0 | 0 | 0 | **1** |
| North West of England | 2 | 3 | 1 | 0 | **6** |
| South East of England | 1 | 2 | 3 | 0 | **6** |
| South West of England | 4 | 0 | 4 | 1 | **9** |
| West Midlands | 5 | 2 | 1 | 1 | **9** |
| East Midlands | 0 | 1 | 1 | 1<sup>g</sup> | **3** |
| **Ethnicity** |  |  |  |  |  |
| White | N/A | 19 | 12 | N/A | **31** |
| Other | N/A | 2 | 0 | N/A | 2 |
| Not specified | N/A | 1 | 0 | N/A | 1 |
| **Who coordinates care?** |  |  |  |  |  |
| Patient/carer | N/A | 17 | 10 | N/A | 27 |
| GP | N/A | 1 | 0 | N/A | 1 |
| Member of health care team | N/A | 1 | 0 | N/A | 1 |
| GP and patient/carer | N/A | 2 | 1 | N/A | 3 |
| Other | N/A | 1 | 0 | N/A | 1 |
| Don't know | N/A | 0 | 1 | N/A | 1 |

N/A = not applicable as patients/carers and health care professionals were asked different eligibility questions.
a Initially had 23 participants but 1 withdrew their data post focus group
b Two of the interview participants also took part in the workshops
c A range of health care professionals were included within our sample including consultants from various specialities, specialist nurses, GPs, allied health professionals (speech and language therapists, physiotherapists, occupational therapists), genetic counsellors, pharmacists, coordinators.
d Charity representatives were from a range of charities which represented patients with rare conditions
e Some of the participants had multiple roles within the professional category, e.g. being a health care professional and a commissioner, or being a health care professional and a charity representative.
f We asked participants if they attended a specialist service or not. Responses may include seeing specialists in their condition in addition to specialist services.
g Role covers both locations
h A few health care professionals/charity representatives also had personal experience of rare conditions as patients/carers

### Taxonomy of care coordination for rare conditions

Our final taxonomy of care coordination consists of six domains: (1) ways of organising care; (2) ways of organising the professionals involved in a patient’s care; (3) responsibility for coordination; (4) how often appointments and coordination take place; (5) access, and (6) mode (see Table 4). Each domain outlines different ways of coordinating care (labelled ‘sub-domains’). Within each way of coordinating care there are different options (labelled ‘options’).

**Table 4.**
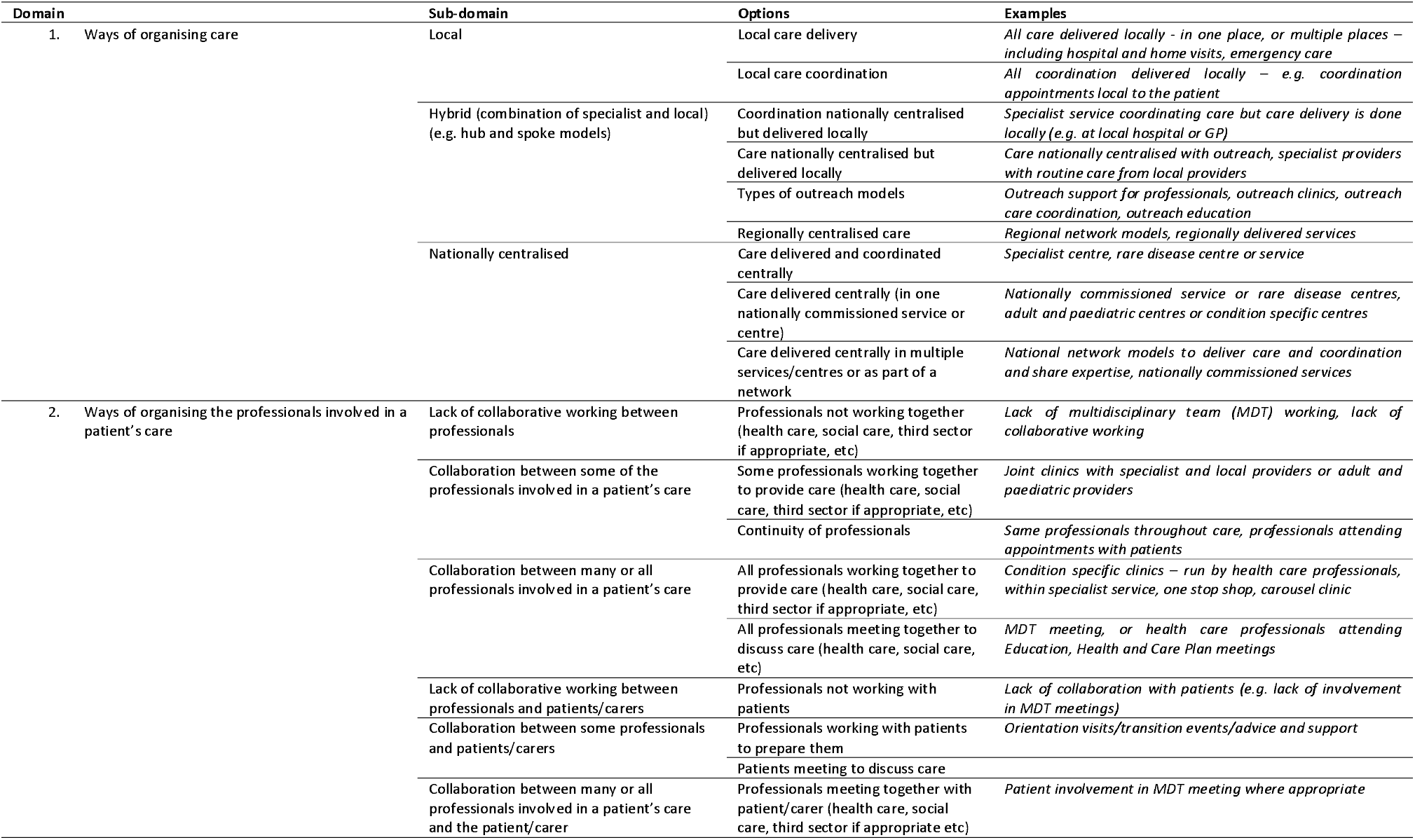
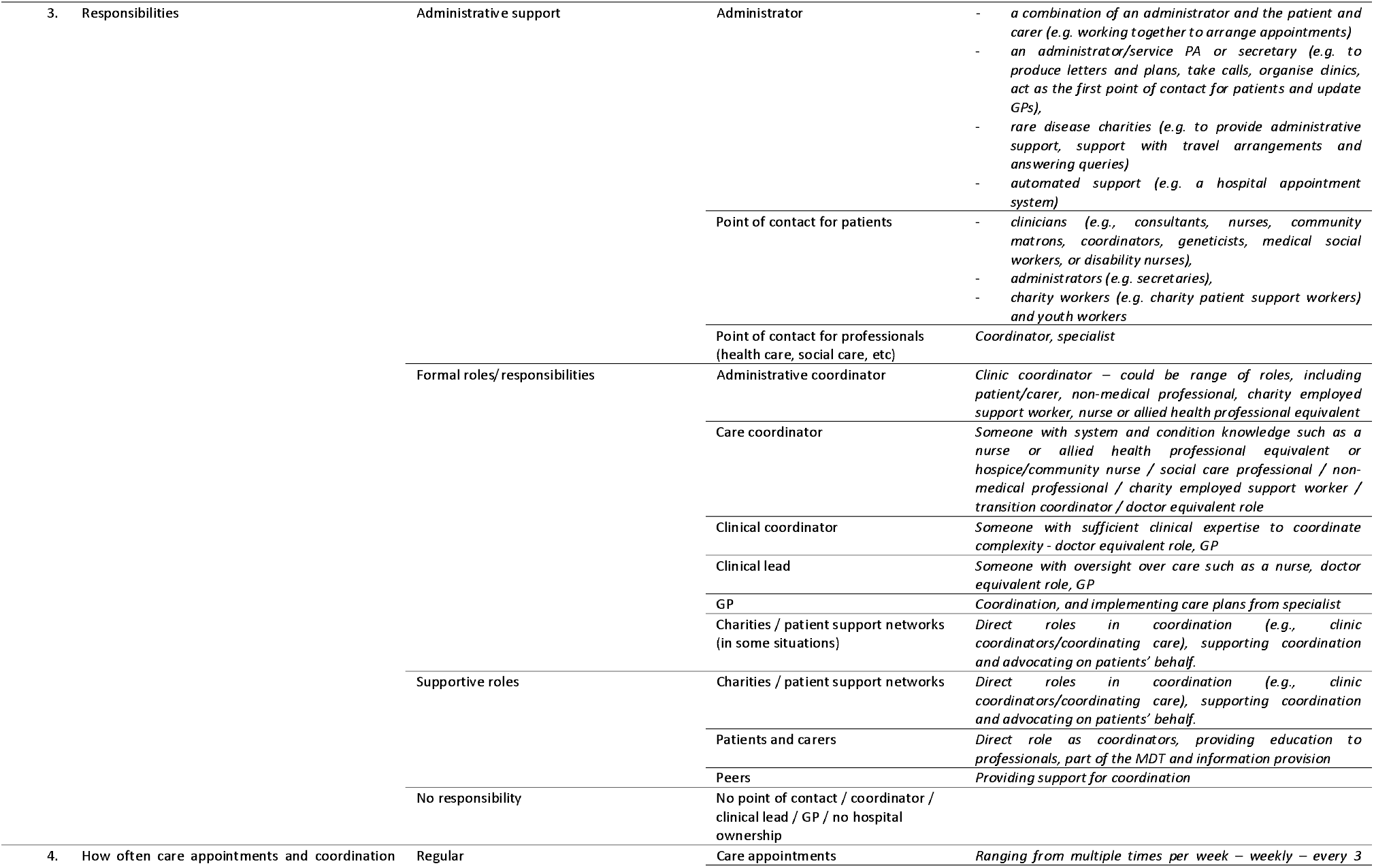
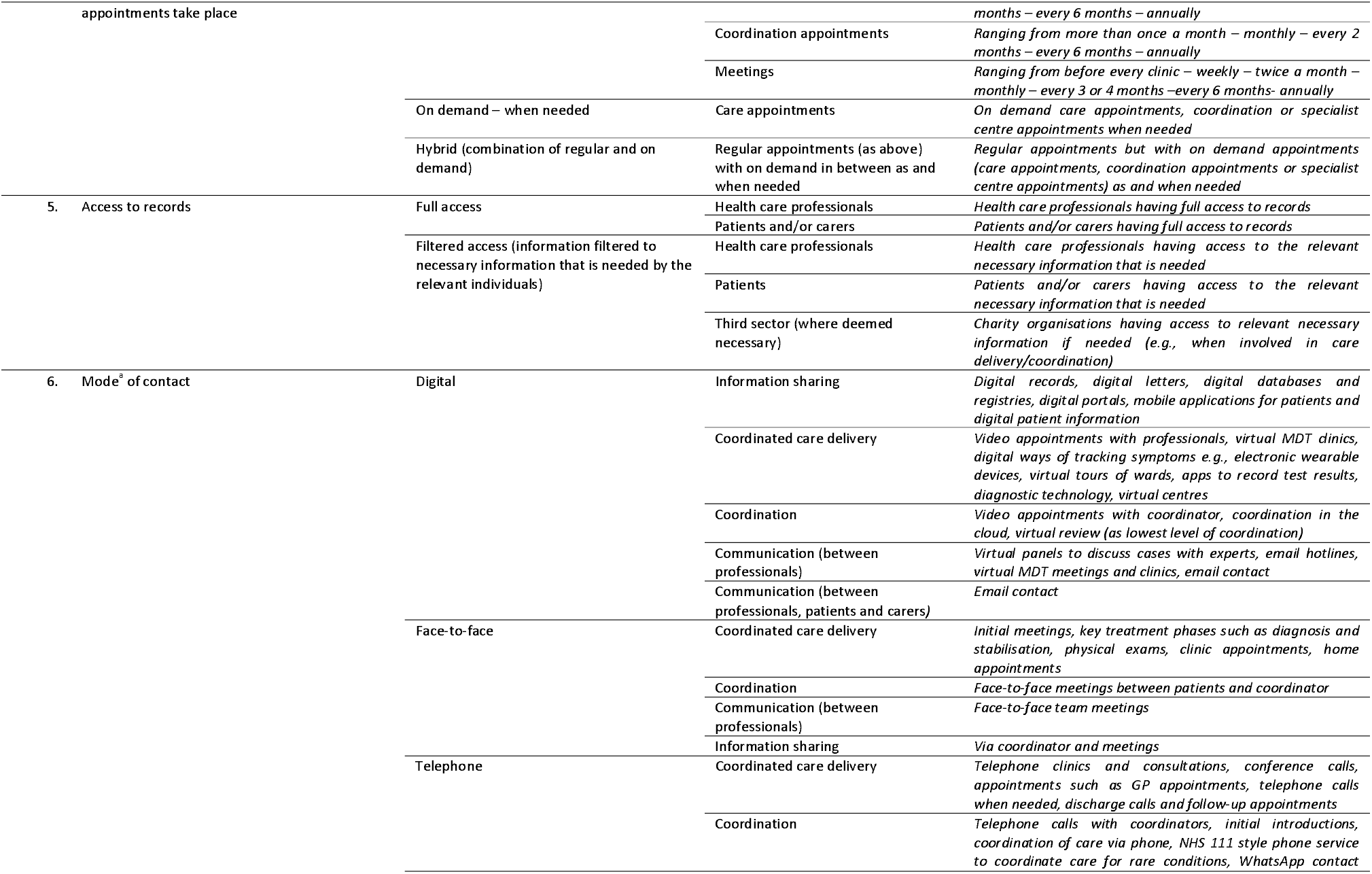
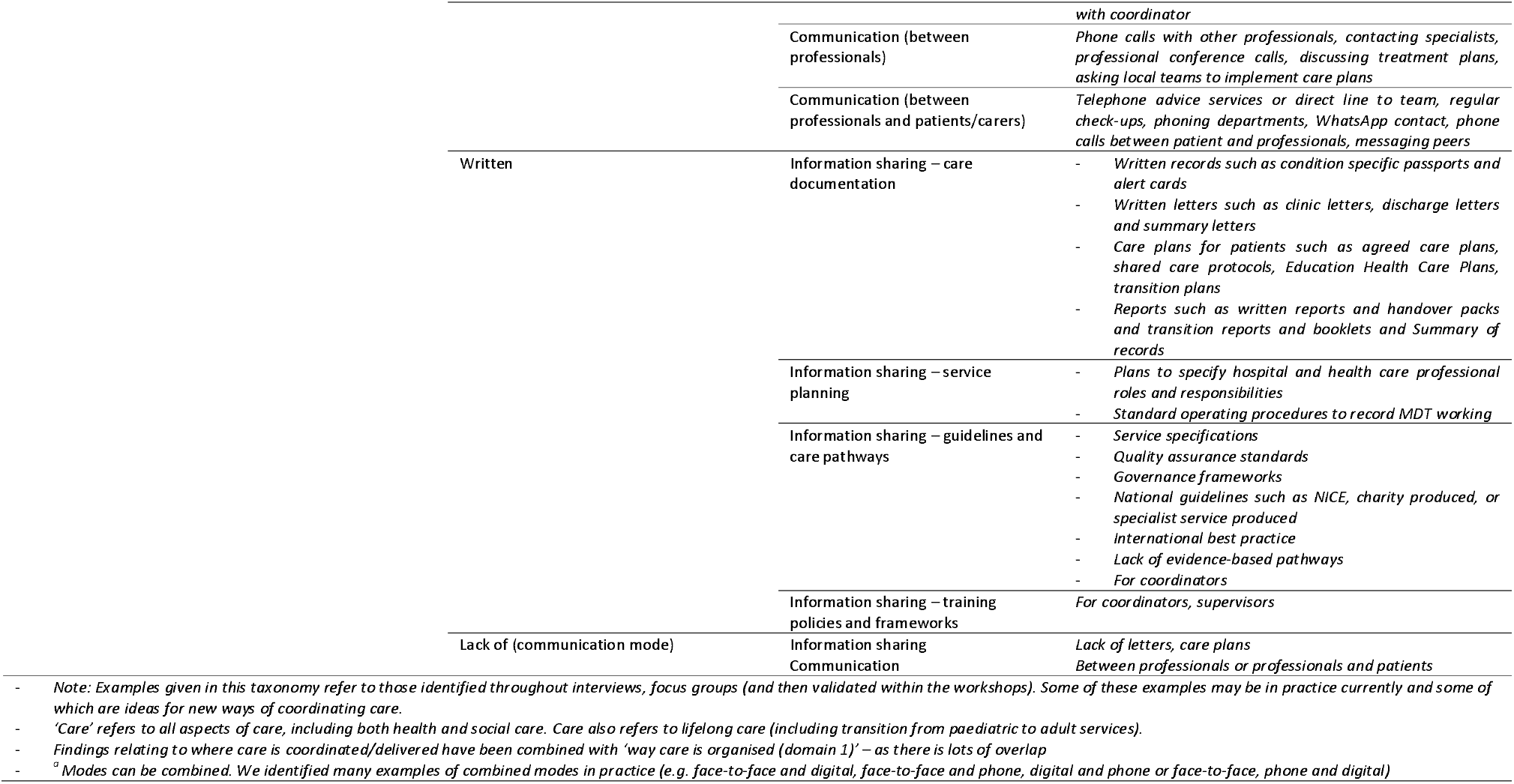
Taxonomy of care coordination for rare conditions

Within the next section we will present each of the taxonomy domains and their sub-domains in turn. Example quotes for each of the six domains are shown in Table 5.

**Table 5.**
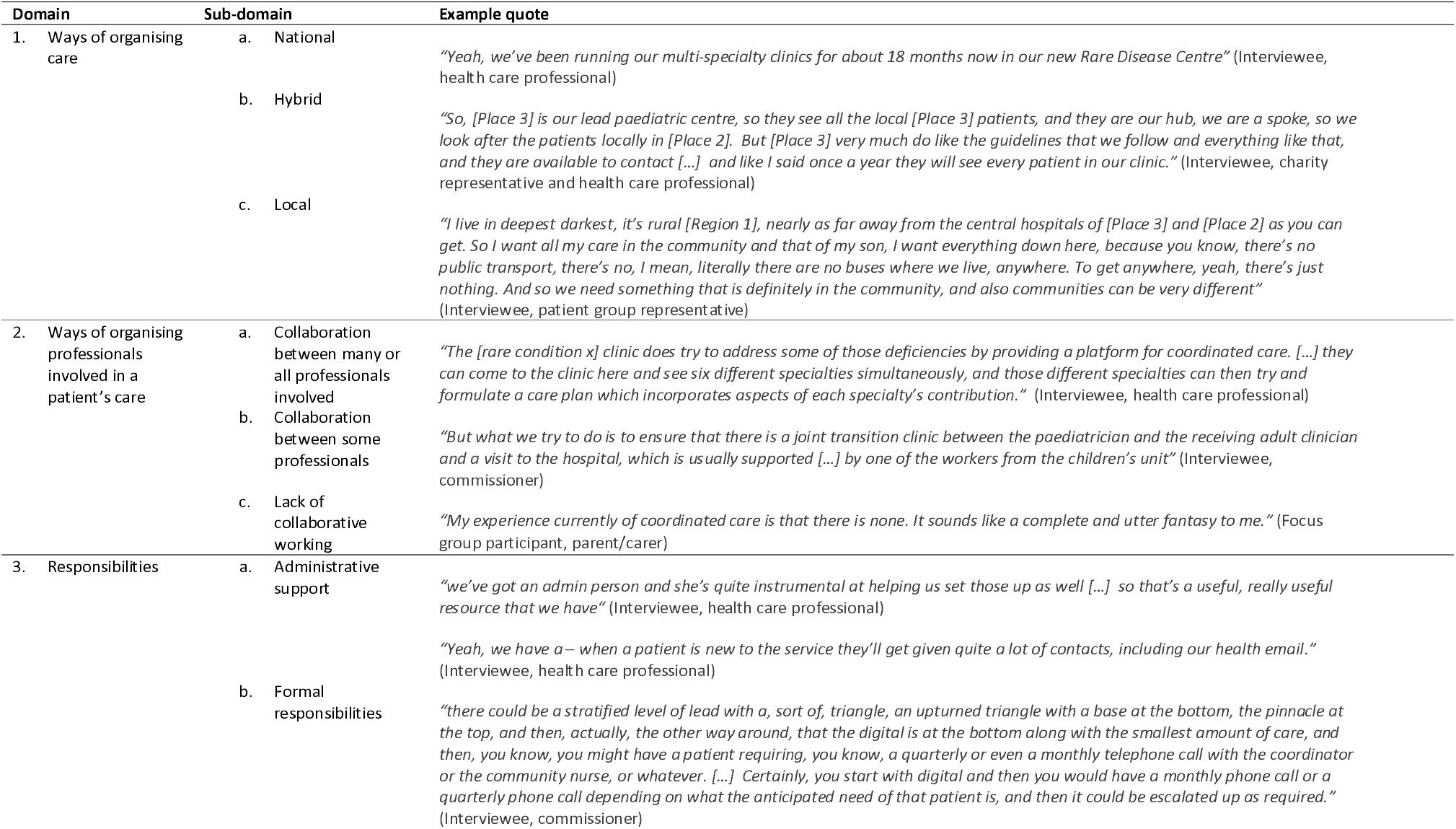
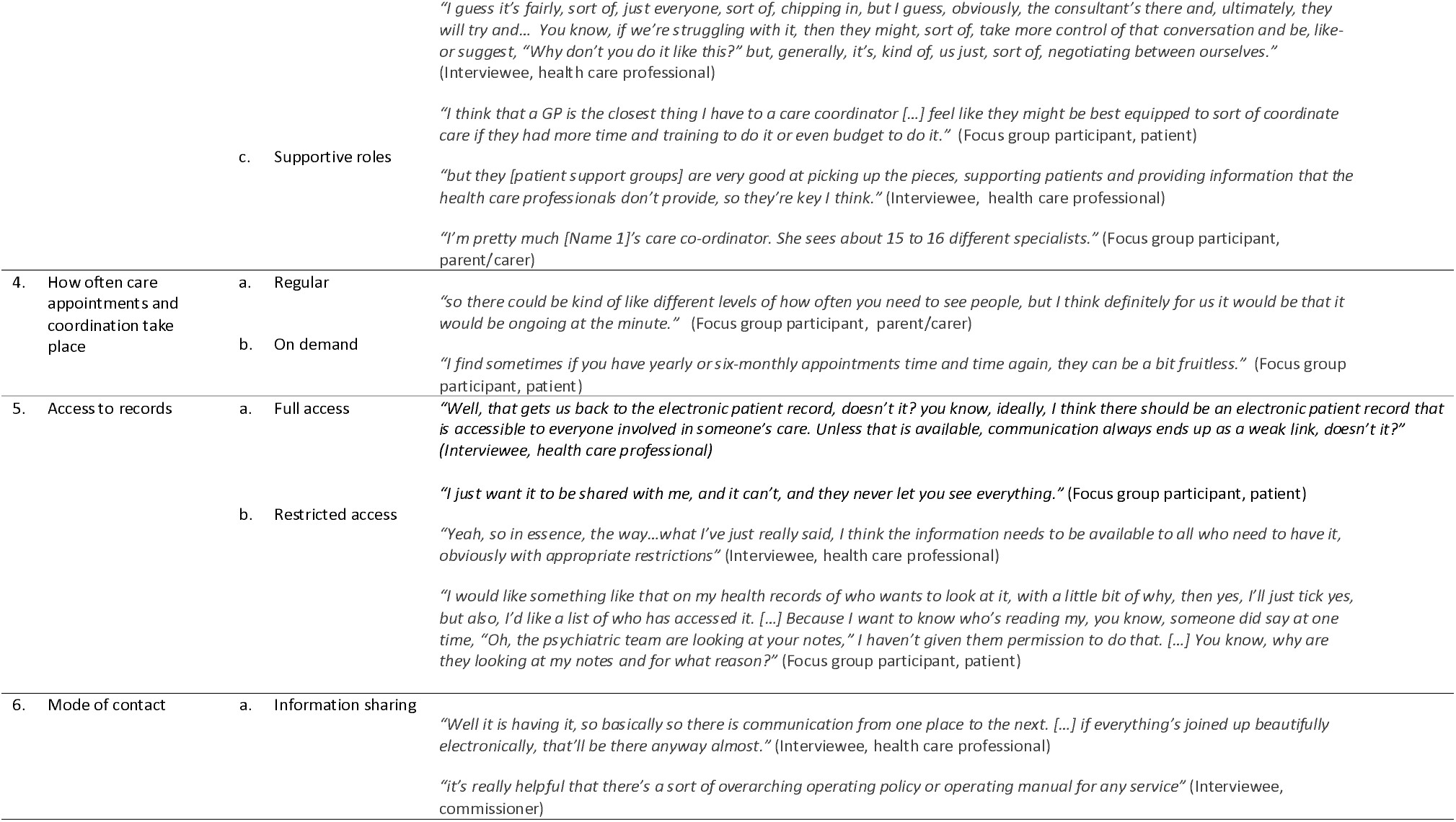
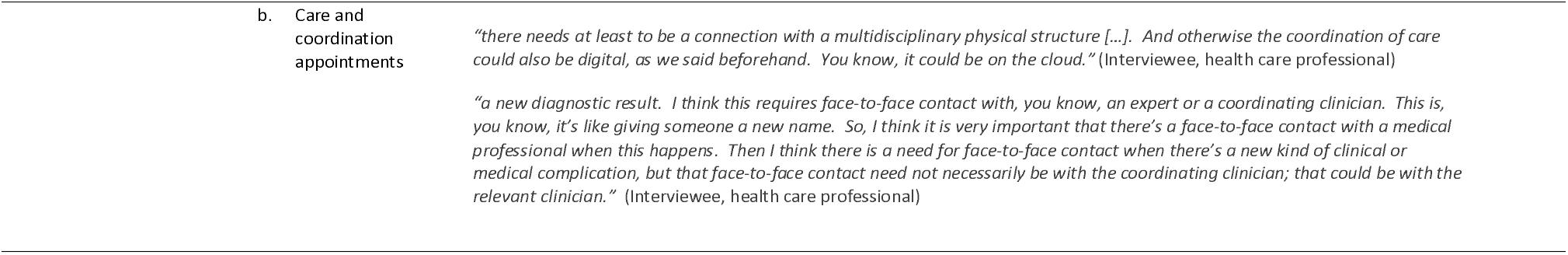
Example quotes (from interviews and focus groups) for each of the six domains.

### Domain 1. Ways of organising care

Our findings highlighted different ways of organising care. Options ranged from local care provision where all care is delivered locally, through to care being delivered in a single national centre that serves all patients in the country with a particular rare condition. Figure 2 provides a summary of the different ways of organising care.

**Figure 2.**
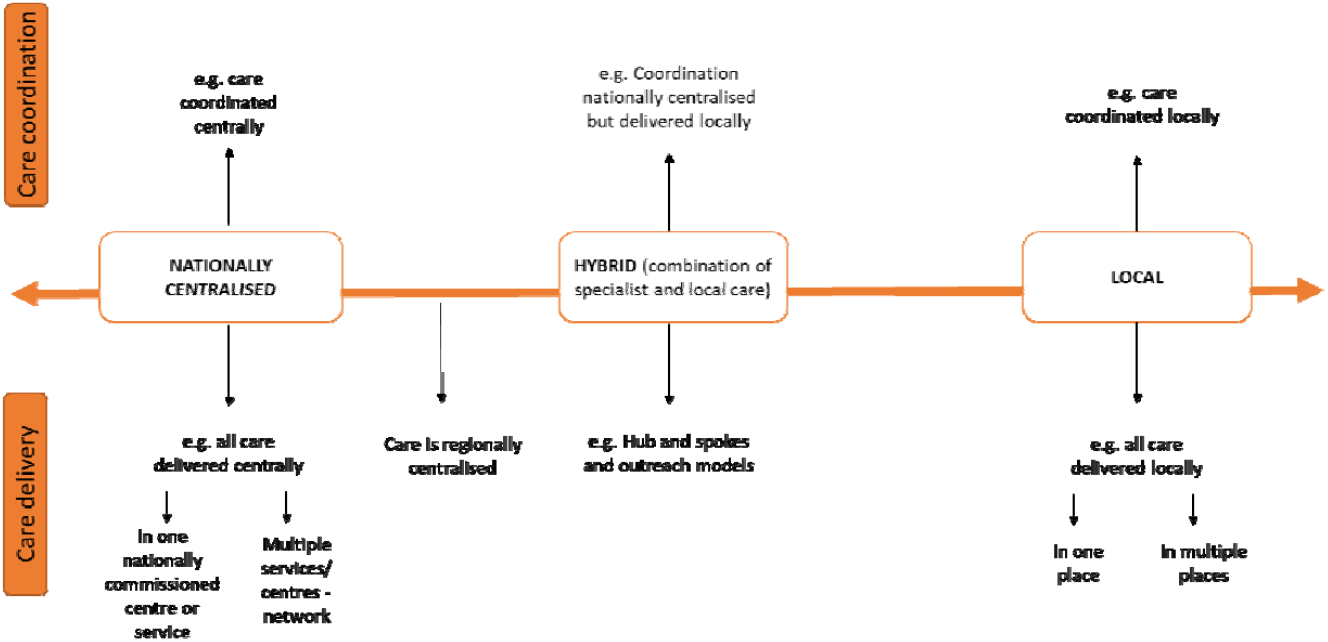
Ways of organising care (visual representation of taxonomy domain 1)

#### Nationally centralised

For nationally centralised services, we identified different examples of nationally centralised care where care is delivered or coordinated centrally. Central delivery of care can either take place in one nationally commissioned centre/service (such as rare disease centres or condition-specific centres) or a network of multiple services or centres.

#### Hybrid

We also identified some ‘hybrid ‘options, which combine both national or regional specialist and local care. Hybrid options include hub and spoke networks, and outreach. There are different types of hub and spoke models. For example, in one type of hub and spoke model of care, the national centre or centres (hub) coordinate care but the actual care delivery happens at a local hospital or GP (spokes). In other types of hub and spoke models, the national centre (hub) provides some care delivery, but other aspects of care are delivered at local hospitals or GPs (spokes). There are also different types of outreach models. Examples of outreach models include outreach clinics (e.g. local outreach clinic, specialists travelling to provide joint clinics with local team, specialists providing care locally) and outreach relating to care coordination (e.g. outreach model of clinical case management in mental health practice, coordinator doing outreach work with local providers and GP and coordinator travelling to provide care locally). Outreach models relating to education included specialist teams providing support for local teams (e.g., education to raise awareness, providing guidance and supervision, email hotline, training, opportunities for local providers to shadow clinics, and formalised agreements that specialists will answer GP queries).

#### Local

Findings highlighted the importance of specialists being involved in care for rare conditions. However, findings also indicated that routine care and non-standardised or tailored care should be delivered locally, and that regular contact with local professionals would be useful. On the other hand, some focus group participants reported wanting all of their care to be delivered locally, or regionally. For some, there was a lack of local care provided.

### Domain 2. Ways of organising the professionals involved in a patient’s care

Our findings highlighted different ways of organising professionals involved in a patient’s care. Options ranged from collaboration between many or all professionals involved in a patient’s care, to collaboration between some of the professionals involved in a patient’s care, to a lack of collaborative working (See Table 4 for examples). Workshop findings highlighted that COVID-19 may have offered new opportunities for collaboration, such as the ability for local team members to dial into multidisciplinary team meetings. Figure 3 provides a summary of the different ways of organising teams.

**Figure 3.**
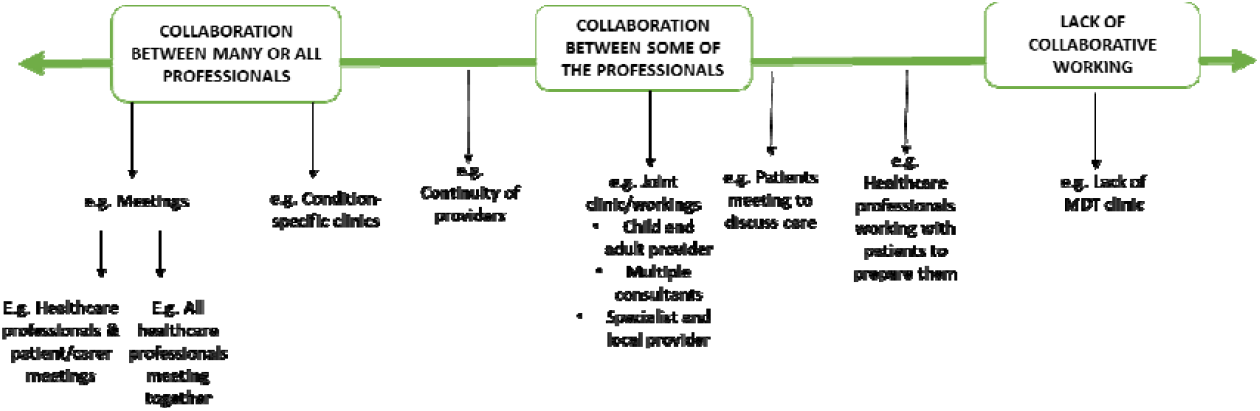
Ways of organising teams (visual representation of taxonomy domain 2)

#### Collaboration between many or all of the professionals involved in a patient’s care

Examples of collaboration between many or all of the professionals involved in a patient’s care includes condition specific clinics (for example those organised and led by individual health care professionals and those delivered as rare disease or specialist clinics) and multidisciplinary meetings held between all professionals (and the patient/carer where appropriate). Different types of condition-specific clinics exist, ranging from: multidisciplinary team appointments including all professionals; one stop shops where patients receive all care in one place at the same time; multidisciplinary clinics that involve professionals seeing patients both together as a team but also separately; and carousel clinics whereby the health care professional moves around whilst the patient stays in the same room.

#### Collaboration between some of the professionals involved in a patient’s care

One example of collaboration between some of the professionals involved in a patient’s care is joint clinics. Joint clinics consist of a couple of professionals working together to provide care. For example, joint clinics consisting of an adult and a child provider; joint clinics consisting of multiple consultants; and joint clinics consisting of specialist and local providers. Additionally, close working between different professionals may occur (e.g. paediatrician contacting adult provider when the patient is ready to transition to adult care).

#### Lack of collaborative working

In some cases, examples demonstrating a lack of collaborative working between professionals involved in a patient’s care were identified, including a lack of multidisciplinary team clinic, lack of transition methods and lack of ownership.

### Domain 3. Types of responsibilities and roles needed for care coordination

Our findings highlighted different types of responsibility and roles involved in coordinating care for rare conditions (administrative, formal and supportive roles) across health care, social care and voluntary sectors. Figure 4 provides a summary of the different types of responsibility and roles.

**Figure 4.**
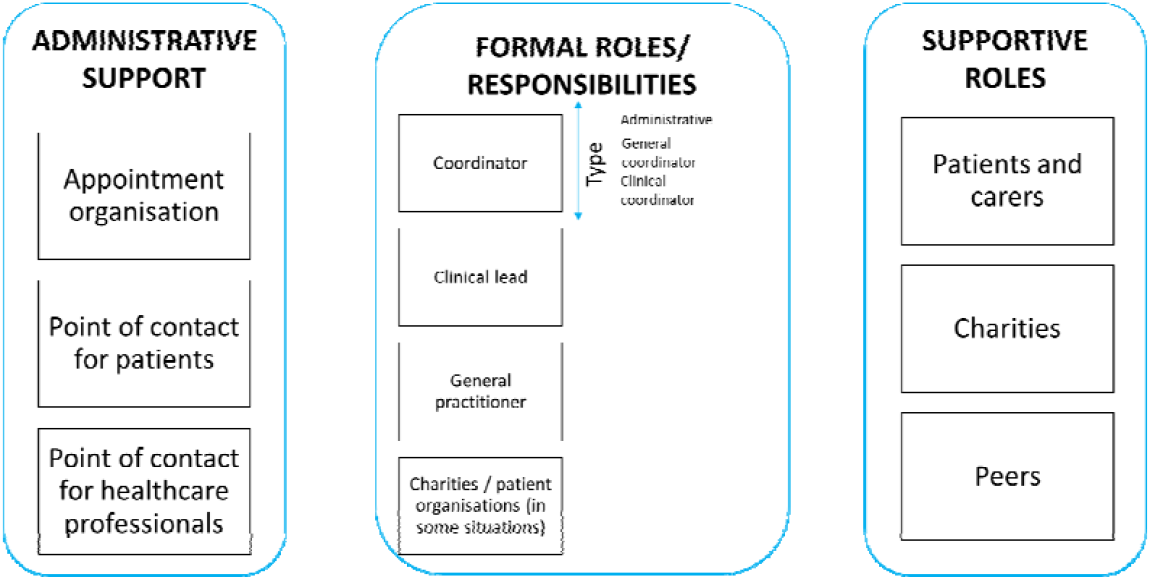
Types of responsibilities for coordination (visual representation of taxonomy domain 3)

#### Administrative support

Administrative support for appointment organisation was highlighted as important. Additionally, having a point of contact for patients and having a point of contact for other professionals was felt to be important. Some patients and carers highlighted that they do not currently have a point of contact. Different options were highlighted in relation to who should provide administrative support (e.g. administrator and patient/carer, administrator, charities, automated support) and who should be the point of contact for patients (e.g. clinicians, administrators, charity workers).

#### Formal coordination responsibilities

Formal coordination responsibilities across three roles were identified: i) those conducted by a coordinator, ii) those conducted by a clinical lead in secondary/tertiary care, and iii) those conducted by a GP in primary care.

Our findings highlighted many roles of a care coordinator (e.g., liaising with health care professionals, coordinating the MDT and aspects of care across different sectors, trusted named person for the patient, ownership, quality assurance and planning). Many different types of coordinators were identified: i) administrative coordinators, ii) care coordinators and iii) clinical coordinators. Administrative coordinators are individuals who arrange MDTs and clinics (e.g. patient/carers, non-medical professionals, charity employed social workers, nurse or allied health professionals). Care coordinators are individuals who have a formal/professional role for coordinating care in addition to system and condition knowledge (e.g. specialist nurses, allied health professionals, hospice/community nurses, social care professionals, non-medical professionals, charity employed social workers and transition coordinators). Clinical coordinators are individuals with sufficient clinical expertise to coordinate complex cases (e.g. doctors or GPs).

Our findings highlighted many roles of a clinical lead, including overseeing or managing care in a service, clinical case management, supervision of professionals, decision-making about extent to which different levels of coordinator are needed, and delegating and ensuring accountability of responsibilities. Clinical leads were identified from a range of roles (e.g. consultants, discipline-specific clinical leads, paediatricians (e.g., hospital/community), the patient’s favourite doctor, a specialist nurse, or geneticists). Some patients and carers reported that they did not have a clinical lead responsible for their care.

Findings highlighted that GPs may have the potential to be involved in coordination in numerous ways, including as a coordinator (e.g. making appointments, named coordinator, developing care plans, identifying when patients need to see coordinator), clinical lead (e.g. a role between primary care and tertiary care to enable them to be responsible for care, or having a GP equivalent role for rare conditions), and implementing care plans provided by specialists (e.g. gatekeeper for specialist care referrals, providing local care and implementing care plans). However, findings indicate that many GPs do not take ownership and that some patients do not have a named GP. A lack of communication between GPs and specialists indicated a need for further collaboration between GPs and specialists (e.g., involvement in MDTs, working in hospital settings, receiving training by specialists, formalised contracts, point of contact).

#### Supportive roles

Supportive roles were also identified, including those conducted by charities/patient organisations and carers and those conducted by patients/carers (see Table 4).

Findings indicated that charities/patient organisations play many roles in coordination including supporting coordination by providing support to patients/carers (e.g. providing information, holding support groups, providing helplines) and professionals (e.g. training of professionals and guiding coordinators). Charities also have direct roles in coordination (e.g. funding coordinator posts and clinical networks, being clinic coordinators, coordinating care, providing resources to help coordinate care, and managing registries). Whilst charities provide funding currently there were views that charities should not be filling gaps for health care services and that charities may not have the capacity to be the main coordinator. Additionally, charities play a role in service quality and improvement (e.g., care pathway development, pulling together evidence, identifying weaknesses in coordination, and setting up/developing specialist services).

Findings indicated that patients and carers currently have lots of direct involvement in coordinating care. Patients and carers act as coordinators of care (e.g. coordinating care across multiple hospitals, being their own advocates, taking more responsibility and chasing appointments) or collaborating with health care professionals to coordinate care (e.g. to arrange appointments, support from health care professionals for coordination as and when needed, and wanting care to be coordinated in partnership with themselves). Patients and carers also support transition to adult services, provide education to health care professionals, and monitor their own care.

### Domain 4. How often care appointments and coordination take place

Our findings highlighted different time periods for care appointments and coordination activities. Options included regular appointments, on demand appointments and hybrid of regular and on demand appointments (see Table 3). Workshop findings highlighted that COVID-19 may have provided more opportunities for on demand appointments for those with stable conditions. See Figure 5 for a summary of this domain.

**Figure 5.**
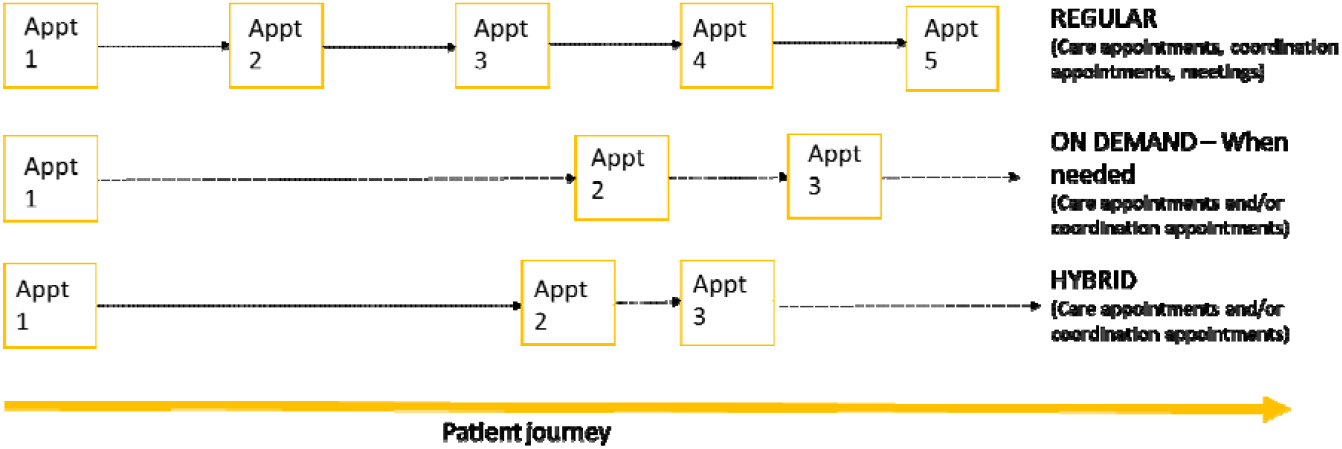
Different options for how often care appointments (specialist and non-specialist) and coordination appointments take place (visual representation of taxonomy domain 4)

#### Regular

Regular appointments were discussed in relation to: frequency of regular specialist centre appointments (ranging from six weeks post treatment to every 18 months), frequency of regular care appointments (ranging from multiple appointments in one week to every 6 months), frequency of contact (ranging from monthly check-ins to yearly check ins), frequency of outreach visits (e.g. annually), frequency of contact with coordinator (ranging from monthly to annually) and frequency of MDT meetings between health care professionals (ranging from weekly to twice a year). Participants indicated that the regularity of appointments is and should be guided by condition-specific guidelines (where available).

#### On demand

Findings from some focus group participants and some interviewees indicated that on demand contact or care appointments (with specialists and coordinators) may be better than regular contact for some patients. One caveat to on demand appointments was that there needs to be quick access to expertise and care in emergencies.

#### Hybrid

Workshop findings highlighted the need for a hybrid category that combines both regular care (at a minimum) with on demand support.

### Domain 5. Access to records

Our findings highlighted different types of access. Options ranged from full or restricted access to records for patients and providers and access to support for patients and health care professionals (see Table 4).

For patients, findings highlighted the need for patients and carers to have access to their own information. This included: access to their records, and access to meetings concerning them.

For health care professionals, findings indicated the importance of access to information and records, given that patients are often seen in different places and by different professionals. The extent to which health care professionals should access information and records was contested; with some patients and carers thinking that any health care professional involved in care should be able to access records, and others thinking that this access should be limited (e.g., to necessary information such as the information relevant to the current care/condition, rather than all of the patient’s history). Some patients highlighted that they would like to control who has access to this information. Workshop participants highlighted that full access to records with a summary may be helpful.

### Domain 6. Mode of contact

Our findings highlighted different modes (see Table 4). Figure 6 provides a summary of this domain.

**Figure 6.**
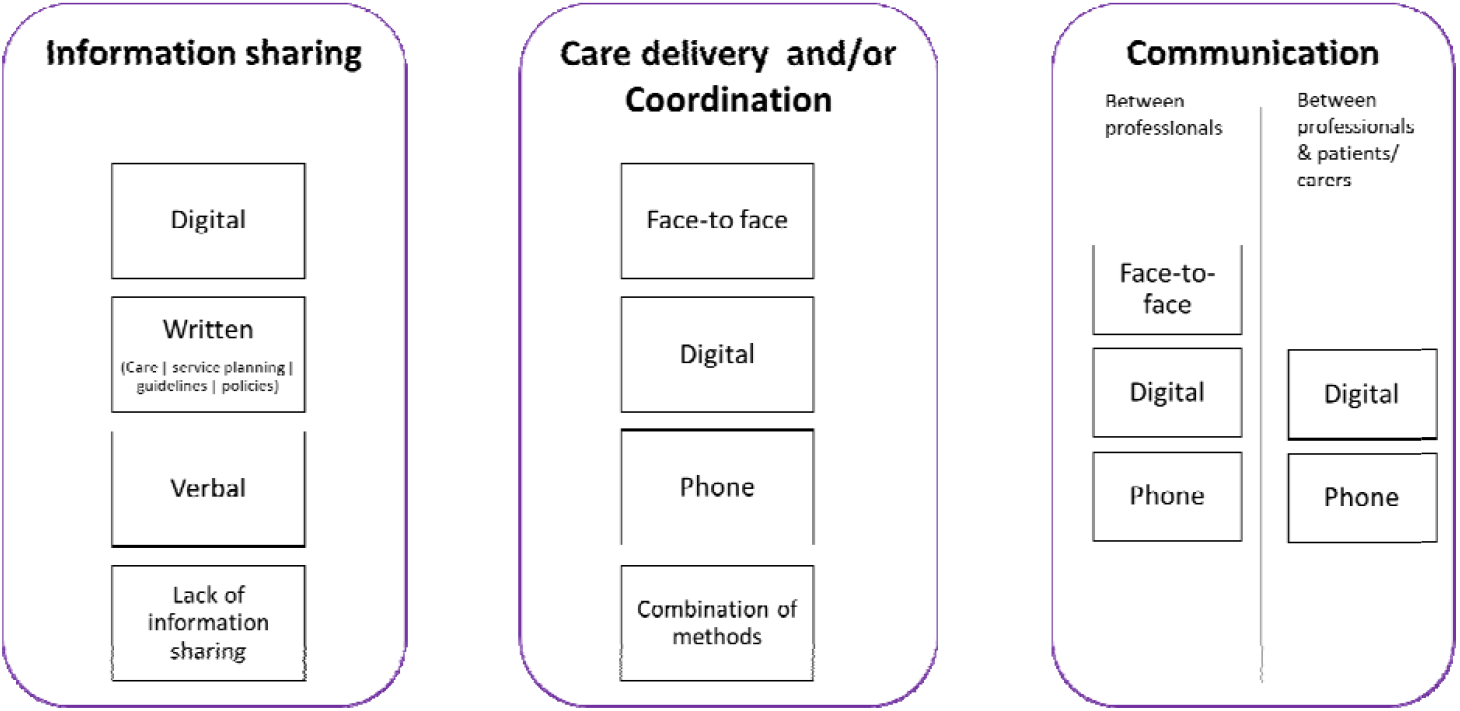
Different options for mode of coordination (visual representation of taxonomy domain 6)

#### Information sharing/communication

For information sharing, participants described many different modes, including digital methods, written methods, verbal methods or a lack of information sharing.

Options for digital information sharing included digital records, letters, databases and registries (stored locally, in the cloud or on an app), portals (e.g. national online portals to access records, letters and guidance), mobile applications for patients (e.g. Patient Knows Best app or apps for patients to hold and share information from their records), and patient information. Digital options for communication between professionals (e.g. virtual panels, email hotlines, virtual meetings, email) and between professionals and patients (e.g. email, Whatsapp) were identified.

Many different written methods of information sharing were also identified: i) written care documentation, ii) service planning, ii) guidelines and pathways. For written care documentation, this included: written records (e.g., condition specific passports and alert cards), letters, care plans (e.g. agreed care plans, education health and care (EHC) plans and transition plans) and reports. For service planning, written methods included plans to specify hospital and professional roles and responsibilities, and standard operating procedures to record MDT working. For guidelines and pathways, this included evidence-based service specifications outlining service standards, quality assurance standards, national guidelines (e.g. NICE or charity produced), international best practice and training policies and frameworks for care coordinators and supervisors.

Telephone was also identified as a mode of communication between professionals (e.g. ringing others to coordinate care, conference calls, discussing treatment plans) and between professionals and patients (e.g. emergency point of contact and telephone advice services).

A lack of information sharing was also highlighted throughout the focus groups and interviews (including a lack of communication between team members and a lack of information sharing).

#### Care and coordination appointments

In terms of care and coordination appointments and communication, our participants described many different modes, including face to face, digital, phone and a combination of methods.

Face-to-face care delivery was identified for many aspects of care and coordination, including meetings (e.g., team meetings and patient/coordinator meetings), care appointments (e.g. initial patient/professional contact, at key treatment phases such as diagnosis and stabilisation, and for appointments requiring physical examinations) and support (e.g. peer support group meetings, network member meetings, and monitoring from charity nurses).

Digital options for care delivery and coordination appointments were identified. For coordinated care delivery, this included: video appointments with professionals (e.g., Skype, Zoom, WhatsApp, Facetime), virtual centres and MDT clinics, digital monitoring (e.g., electronic wearable devices, apps to record test results), virtual tours of wards, diagnostic technology. For coordination this included: video appointments with coordinators (e.g., Skype, Zoom, Whatsapp, Facetime), coordination in the cloud, and virtual review (as the lowest level of coordination).

Options for telephone care delivery (e.g., clinics/consultations and catch ups) and coordination (e.g. calls with coordinators and introductions) were also discussed. Workshop findings highlighted that COVID-19 has accelerated the shift to digital and telephone delivery of care for people living with rare conditions.

## Discussion

### Key findings

We have developed a taxonomy of care coordination for rare conditions, based on learning from the UK health system and the National Health Service. We identified six domains of care coordination: (1) ways of organising care (local, hybrid, national); (2) ways of organising the team (collaboration between many or all professionals, collaboration between some professionals a lack of collaborative working); (3) responsibility for coordination (administrative support, formal roles and responsibilities, supportive roles and no responsibility); (4) how often appointments and coordination take place (regular, on demand or hybrid); (5) access (full or filtered access to records), and (6) mode of information sharing, care coordination/delivery and communication.

### How findings relate to previous research

These findings extend knowledge on care coordination. National policy documents and previous research have highlighted the importance of care coordination [3,13,32]. However, findings indicate that little is known about coordination for rare conditions and what this might entail [13]. Previous research has shown that coordination for rare and common chronic conditions has many components [12,13,33,34], but care coordination had not been formally categorised. The taxonomy presented in this paper extends previous research by formalising care coordination for rare conditions into six domains (each with different options). Whilst previous research has developed taxonomies for related concepts [16,17,18,19], this is the first research that has attempted to develop a taxonomy of care coordination for rare conditions. Findings indicate that whilst different conditions have different characteristics and challenges, it is possible to develop one taxonomy that covers a range of conditions and a range of care coordination options.

Our findings highlighted three main options for organising care, including nationally centralised, hybrid and local care. This supports previous research which has highlighted the importance of specialist centres for people living with rare conditions [35] but extends previous research by demonstrating the potential usefulness of hub and spoke models and outreach models for rare conditions; models which are not new but which have been used in other chronic conditions with success [36,37,38]. Additionally, the findings highlight that local care is not necessarily problematic, with some participants articulating the role of local care and the benefits that this provides them. However, findings do indicate that developing local expertise and knowledge is key.

Our findings on the organisation of professionals for rare conditions supports previous research that indicates the importance of collaboration and MDTs for rare conditions [12,39] and other conditions [40]. Findings also support previous research which indicates a need to join up care appointments from different disciplines and hospitals into one appointment (e.g., condition-specific clinics), in order to facilitate coordination [12,13]. However, findings indicate that collaboration does not always happen in practice and that improvements in collaboration/joined-up working are needed. However, there have been some recent initiatives to improve collaboration across health and social care generally (e.g. the introduction of care coordinators in primary care networks).

Our findings extend previous research by outlining different types of responsibilities and roles needed to coordinate care for rare conditions. Previous research indicates the importance of care coordinators [12,13,41,42]. However, findings extend this research by demonstrating the many different roles needed to coordinate care (administrative support, coordinators, clinical leads, GPs, charities and patients/carers). However, patient/carer involvement in coordination is not always appropriate if patients/carers are unable or do not want to coordinate their own care; and may have a negative impact on patients, families and the treatment burden that they experience [2, 12, 19, 39].

Our findings highlight the importance of following clinical guidelines and service specifications which outline how often appointments should take place. However, findings indicate that patient and provider factors need to also be taken into account when considering how often patients should be seen (see [20]).

Findings highlighted the potential for remote methods of coordination, including digital information sharing (e.g., through electronic records), virtual clinics and care coordination appointments. This shift to digital methods has been accelerated during the COVID-19 pandemic. This supports previous research, which indicates that digital methods may show some potential for use in health care delivery [43,44]. Our findings suggest that this may also apply to care coordination, but that remote methods cannot fully replace face-to-face appointments.

### Strengths and Limitations

One strength of our study is that we used robust analysis procedures which strengthen the validity of this study. Twenty percent of data were coded by a second researcher. Additionally, the research team and members of the PPIAG were continually involved in discussions about analysis and findings. We also triangulated findings outputs from other parts of the CONCORD study [13, 21] to ensure no major omissions.

Whilst we sampled from a variety of rare conditions, locations and sectors, we were unable to include every rare condition. Some groups, including individuals from minority ethnic groups and certain roles (e.g. GPs) were under-represented. Therefore, whilst we collected extensive data, and included as many different views as possible, we are unlikely to have captured every possible option of care coordination. However, our PPIAG was a diverse and representative group. Additionally, the taxonomy was developed from data collected within the UK, and therefore it is likely that findings may only apply to the UK healthcare system. However, it is hoped that the taxonomy will also provide learnings in other healthcare contexts.

Previous research has demonstrated that one key challenge of coordination is that it is difficult to distinguish between aspects of care and coordination components [13,45]. This is also a potential limitation of some options within our taxonomy (e.g., mode of care appointments, frequency of care appointments). However, we believe that the mode and frequency of such care appointments is part of care coordination, i.e., it may be that a care appointment that takes place virtually (with all health care professionals involved) may be more coordinated than other modes of care appointment (e.g., visiting different health care professionals for individual appointments). Additionally, this was not identified as a concern by any workshop participants.

### Implications

The development of the taxonomy could lead to the standardisation of terminology for care coordination in rare conditions. Previous research proposed that better measurement of systems for organising and delivering health care systems are needed [1]. This taxonomy will help to achieve this goal as it provides a better understanding of coordination and ways of organising and delivering health care for people affected by rare conditions. This will support researchers in operationalising and measuring care coordination. If care coordination strategies are piloted, evaluated and implemented more widely within the NHS, this may lead to better care and reduced burden for people living with rare conditions [9,46].

The taxonomy can be used by health care professionals delivering care for people with rare conditions and as a menu for policy makers, service planners, researchers and commissioners to consider when developing new and/or existing models of coordination for rare conditions. For example, we have used the taxonomy, together with the qualifier findings to develop some hypothetical models of care coordination that may be applicable in different situations (see [20]).

We have also developed a flow chart that may inform how these findings are used to develop such models (see [20]). These models can be costed and evaluated by researchers and services. These findings could be particularly helpful during the development of the rare disease action plan in response to the new Rare Disease Framework [47]; in which care coordination is identified as a key priority. The taxonomy can also be used by researchers to evaluate models of care coordination. The taxonomy also provides clinicians and patients/carers with expectations about the different ways in which care can be coordinated. Given similarities between common and rare chronic conditions that were highlighted in previous research [13], researchers interested in care coordination for other conditions may be able to adapt the taxonomy for use in other complex chronic conditions. Additionally, the process outlined in this manuscript could be adapted by researchers to develop comprehensive taxonomies to understand and organise other health care services.

### Future research

Future research is needed to explore where different ways of coordinating care have been implemented and to evaluate the implementation, effectiveness, and cost-effectiveness of different models of care coordination for rare conditions in practice. This is important given that it is not yet clear whether coordinated care leads to better outcomes for patients/carers, professionals and organisations. Further research is needed to operationalise care coordination models so that delivery of care coordination can be measured.

## Conclusions

Findings from our qualitative study with key stakeholders (patients, carers, health care professionals, charity representatives and commissioners) provide a thorough taxonomy of care coordination for rare conditions. Our taxonomy can facilitate the development and evaluation of existing and new models of care coordination for people living with rare conditions. The process outlined in this manuscript provides a template that could be adapted to develop taxonomies for other health care services.

## Data Availability

The datasets generated and/or analysed during the current study are not publicly available due to participant confidentiality but are available from the corresponding author on reasonable request.

## List of abbreviations

CONCORD: Coordinated Care of Rare Diseases Study
COVID-19: Coronavirus Disease-19
GP: General practitioner
HS&DR: Health Services and Delivery Research programme
MDT: multidisciplinary team
NHS: National Health Service
NICE: The National Institute for Health and Care Excellence
NIHR: National Institute for Health Research
PPIAG: Public Patient Involvement Advisory Group
UCL: University College London
UK: United Kingdom

## Acknowledgements

Thank you to the wider CONCORD team (Professor Lyn Chitty, Sharon Parkes, and Christine Taylor) for providing support and feedback throughout the study. Thank you to the CONCORD Patient and Public Involvement Advisory Group for providing feedback throughout the project. Thank you to all of our participants who took part in the interviews, focus groups and workshops.

