## Appendix 1 and 2 for "Developing a taxonomy of care coordination for people living with rare conditions: A qualitative study"

Appendix 1. Topic guides for interviews and focus groups

**Interviews**

| **Interview questions** | **Prompts** |
| --- | --- |
| 1. Please tell me a bit about yourself. | - Job role - Experience of rare or ultra-rare diseases - Experience providing or being involved in the provision of coordinated care |
| 2. What types of coordinated care are you aware of? | - Models of coordination - Coordination addressing different types of transitions - How is this model similar to other models of coordinated care? - How is this model different from other models of coordinated care? |
| 3. What are the implications of coordinated care/lack of coordinated care? | - Positive - Negative - Please could you give an example of where things went well or didn’t go well? |
| 4. What type of coordinated care would you like to be delivered? | - What would this involve? - How would this be done? - What would this look like if it were successfully used in practice? - Please could you give an example of where this type of coordination went well or didn’t go well? - Please can you tell me a bit about why? |
| 5. What would be your preferred way for patients and family members to access coordinated care? | - What would this involve? - How would this be done? - What would this look like if it were successfully used in practice? - Please could you give an example of where this went well or didn’t go well? - Please can you tell me a bit about why? |
| 6. What would be your preferred format of coordinated care? | - What would this involve? - How would this be done? - What would this look like if it were successfully used in practice? - Please could you give an example of where this went well or didn’t go well? - Please can you tell me a bit about why? |
| 7. What would be your preference on how often patients receive coordinated care? | - What would this involve? - How would this be done? - Would this differ depending on the type of coordinated care provided? - What would this look like if it were successfully used in practice? - Please could you give an example of where this went well or didn’t go well? - Please can you tell me a bit about why? |
| 8. What would be your preferences on where care coordination is provided? (where appropriate) | - What would this involve? - How would this be done? - What would this look like if it were successfully used in practice? - Please could you give an example of where this went well or didn’t go well? - Please can you tell me a bit about why? |
| 9. What would be your preferences on how information would be shared between healthcare providers, patients and carers and local services? (where appropriate) | - What would this involve? - How would this be done? - What would this look like if it were successfully used in practice? - Please could you give an example of where this went well or didn’t go well? - Please can you tell me a bit about why? |
| 10. What would your preferred method of transition (movement) between services (e.g. child to adult) be for patients? | - What would this involve? - How would this be done? - What would this look like if it were successfully delivered in practice? - Please could you give an example of where this went well or didn’t go well? - Please can you tell me a bit about why? |
| 11. How do you think that patients and carers would like their care to be coordinated? | - Please can you tell me a bit about why? |
| 12. What things need to be taken into account when deciding how best to coordinate care? | - E.g. different conditions / different age ranges - Please can you tell me a bit about why? |
| 13. What factors help to provide coordinated care? | - Locally  - Nationally  - How did/might they help?  - Have you been involved in initiatives to improve coordination previously? If so, what changes helped to improve care coordination? |
| 14. What factors get in the way of providing coordinated care? | - Locally  -Nationally  - How did/might they get in the way?  - How do you overcome these problems? |
| 15. Is there anything else that you would like to say about what we have talked about? | - Is there anything else that you would like to mention? - Are there any important issues that have not been raised? |

**Focus group**

| **Structure** | **Prompts (if needed)** |
| --- | --- |
| 16. Let’s begin. Let’s find out some more about each other by going around the table. Please tell us your name, whether you are a patient or parent/carer and where you are from |  |
| 17. Please tell us about your experiences of coordinated care (approx. 2 mins each) | - E.g. fully coordinated care, some coordinated care, no coordinated care   *<After each person>*   - Which aspects of your care that were coordinated worked well? - Which aspects of your care could be coordinated better? |
| 18. What are the implications of having/not having coordinated care? | - Positive - Negative - Please could you give an example of where things went well or didn’t go well? - E.g. number of clinics people attend/how far they have to travel - E.g. psychological, clinical, medical, social, financial implications |
| 19. **Thinking about the different types of coordinated care, please identify your preferred choices for the following aspects of coordinated care:**     - How would you like your care to be coordinated?     - Which aspects of care would care coordination matter most to you?     - Which aspects of care would care coordination not matter to you?     - How would you like to access coordinated care     - How would you like to communicate with other people involved in care coordination?     - How often would you want to receive coordinated care?     - Where would you like care to be coordinated?     - How would you like information to be shared between healthcare providers, patients and carers, and local services?     - What is your preferred method of transition (or movement) across services? | *<Go through each of the questions one by one and prompt the following questions>:*   - Please can you tell me a bit about why this is your preference? - What are other people’s views on this? - How could this be done? - What would this look like if it were successfully used in practice? - Would your preferences change over time? Why? |
| 20. What factors affect your access to coordinated care? | - Locally - Nationally   - How did they help?  - How did they get in the way? |
| 7. What choice do you have in terms of the care coordination that you receive? | - How do you find this? - What choices would you like to make in relation to care coordination? - What could be improved? |
| 8. Is there anything else that you would like to say about what we have talked about? | - Is there anything else that you would like to mention? - Are there any important issues that have not been raised? |

Appendix 2. Topic guide for workshop

| Time | Tasks/sessions |
| --- | --- |
| 10 minutes | Introduction to workshop and ground rules & brief intro to participants & brief recap of video/introduce task |
| 40 minutes | Group discussion on taxonomy (domains and characteristics) – go through each domain answering the following questions:   - What’s good about this domain and the characteristics within it? (10 mins) - What needs improving? (10 mins) - Appropriateness of characteristics within this domain in relation to use during current COVID situation? (10 mins) - Recommendations to improve domain/characteristics? (10 mins)   If time left – could also ask similar questions about the models |
| 10 minutes | Development of recommendations to improve taxonomy and models (summary from discussion and any other thoughts?) |
| 5 minutes | Introduce optional activity for after workshop (if they would like to they can provide feedback on models using the following questions:   - What’s good about the model? - What needs improving? - Appropriateness of model in relation to use during current COVID situation? - Recommendations to improve model?) |
| 5 minutes | Questions and summary/debrief |
