## Supplementary figures and images for "Developing a taxonomy of care coordination for people living with rare conditions: A qualitative study"

### Appendix 3

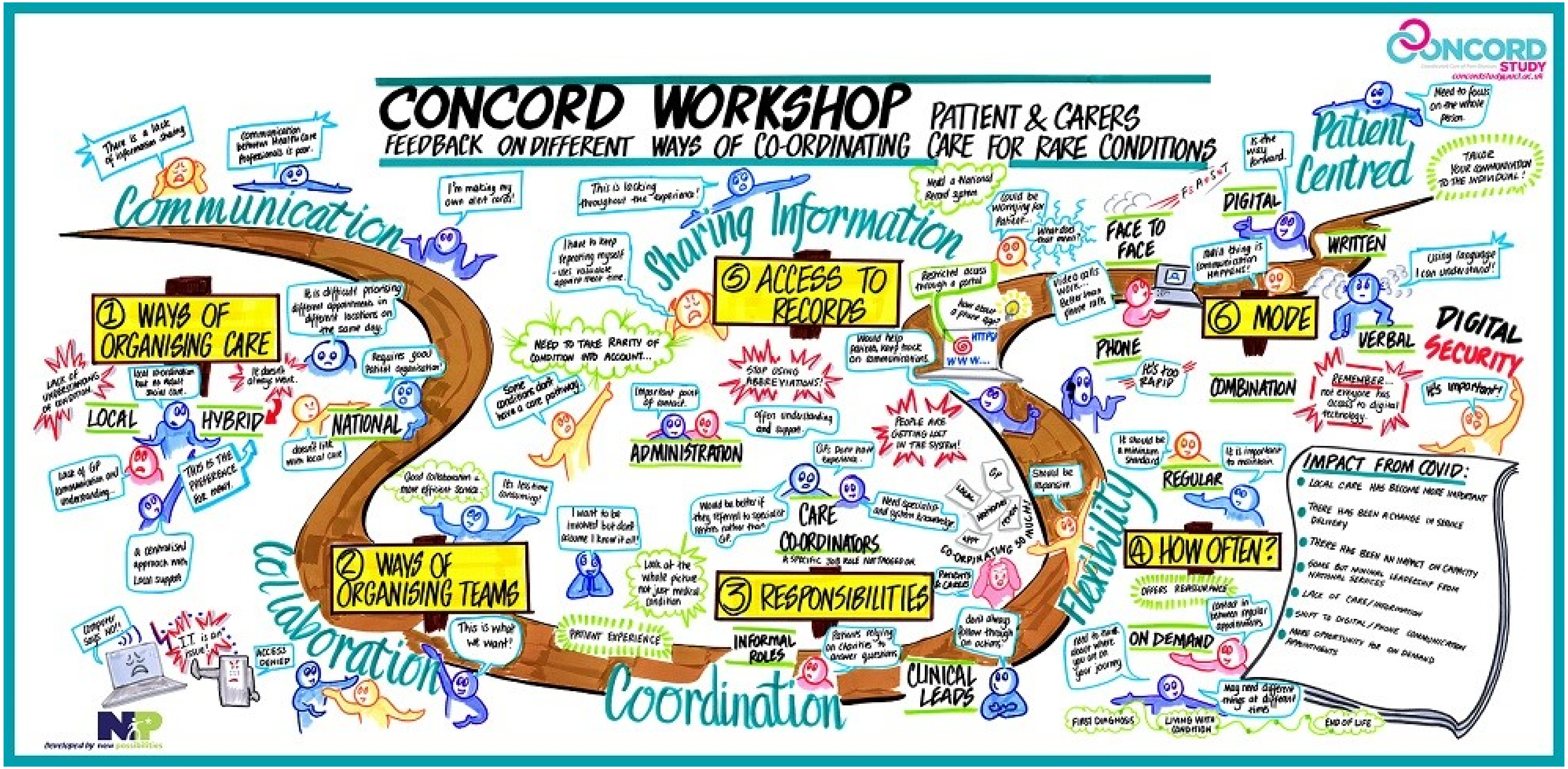

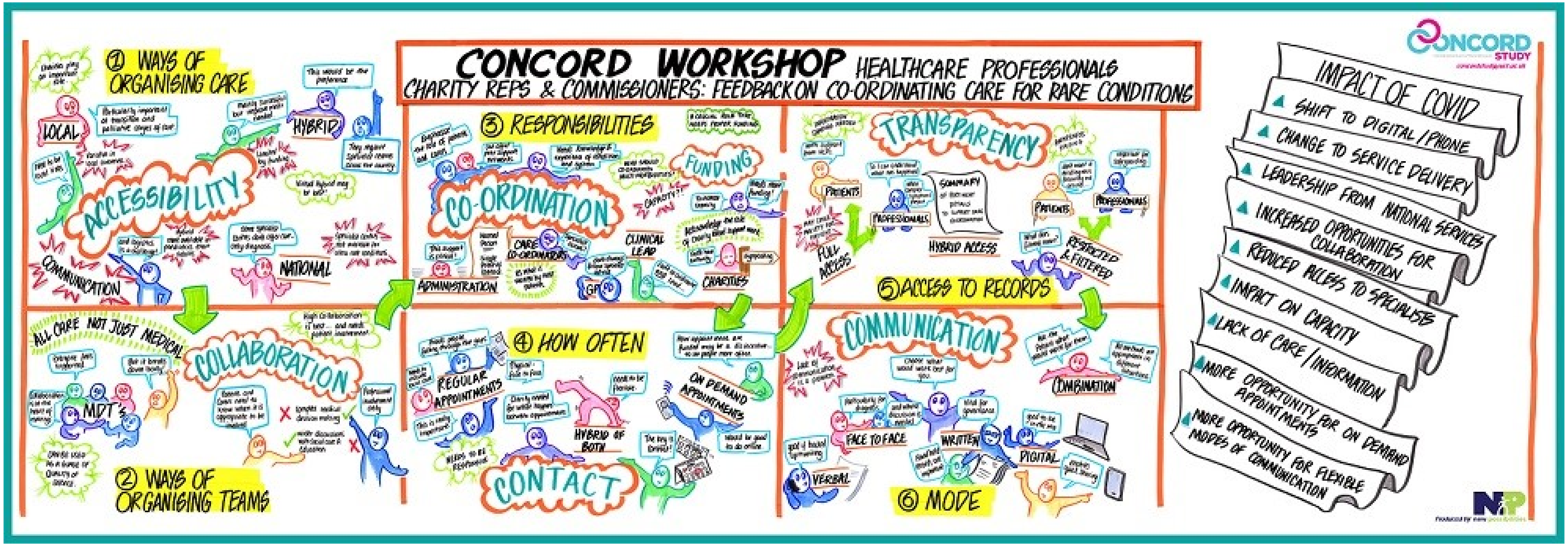
